# Effectiveness of mRNA COVID-19 Booster Vaccines against Omicron and Delta Variants among US Veterans

**DOI:** 10.1101/2022.01.15.22269360

**Authors:** Yinong Young-Xu, Gabrielle M. Zwain, Hector S. Izurieta, Caroline Korves, Ethan I. Powell, Jeremy Smith, Abirami S. Balajee, Mark Holodniy, David O. Beenhouwer, Maria C. Rodriguez-Barradas, Sheldon T. Brown, Vincent C. Marconi

## Abstract

**Background:** We estimated vaccine effectiveness (VE) of mRNA vaccines among US Veterans during periods of Delta and Omicron variant dominance. Patients included in this study were largely 65 years or older (62,834, 55%), male (101,259, 88%), and non-Hispanic white (66,986, 58%).

**Methods:** We used SARS-CoV-2 laboratory test results to conduct a matched test-negative case-control study to estimate VE of three and two doses of mRNA vaccines against infection (regardless of symptoms), and a matched case-control study to estimate VE against COVID-19-related hospitalization and death. We estimated VE as (1-odds ratio) x 100%. Severity of disease was measured using hospital length of stay (LOS) and admission to an intensive care unit (ICU).

**Results:** Against infection, booster doses had 7-times higher VE - 59% (95% confidence interval [CI], 57 to 61) - than 2-dose VE (7%; 95% CI, 3 to 10) during the Omicron period. For the Delta period, estimated VE against infection was 90% (95% CI, 88 to 92) among boosted vaccinees, 64% higher than VE among 2-dose vaccinees [55% (95% CI, 51 to 58)]. Against hospitalization, booster dose VE was 87% (95% CI, 80 to 91) during Omicron and 95% (95% CI, 91 to 97) during Delta; the 2-dose VE was 44% (95% CI, 26 to 58) during Omicron and 75% (95% CI, 70 to 80) during Delta. Against death, estimated VE with a booster dose was 94% (95% CI, 85 to 98) during Omicron and 96% (95% CI, 88 to 99) during Delta, while the 2-dose VE was 75% (95% CI, 52 to 87) during Omicron and 93% (95% CI, 85 to 97) during Delta. During the Omicron period, average hospital LOS was 4 days shorter [3 days (95%CI, 3 to 4 days)] than during the Delta period.

**Conclusions:** A mRNA vaccine booster is more effective against infection, hospitalization, and death than 2-dose vaccination among an older male population with comorbidities.

## INTRODUCTION

We previously reported^1,2^ the effectiveness of COVID-19 mRNA vaccines against infection, hospitalization, and mortality among Veterans Health Administration (VHA)-enrolled Veterans through September 2021, when the major circulating variants in the United States included Alpha, Beta, and Delta and 2-doses of mRNA vaccine were highly effective against any infection, hospitalization, and death. With the rise of the Omicron variant in December 2021-January 2022, we updated our estimates of vaccine effectiveness (VE) to measure continued effectiveness of mRNA vaccines in a population including the entire U.S.

In September 2021, the Centers for Disease Control and Prevention (CDC) Advisory Committee on Immunization Practices recommended a booster dose of COVID-19 vaccines for everyone ≥65 years and those ≥18 years with certain underlying medical conditions.^5,6^ By the time of our study period, the recommendation was expanded to include everyone 18 and older.^7,8,9^ As of February 4^th^ 2022, the VHA had over 4.4 million fully vaccinated Veterans (2-dose of mRNA or 1-dose of Johnson & Johnson’s Janssen COVID-19 vaccine) and over 1.4 million Veterans had received an mRNA booster.^10^ As of the end of January 2022, the VHA reported more than 587,000 COVID-19 cases and 19,600 confirmed deaths.^10^ Early reports^11,12,13,14,15^ showed varying VEs during various waves of the pandemic corresponding to distinct periods of variant dominance.

We sought to estimate VE against infection, hospitalization, and mortality for mRNA vaccines authorized in the U.S. (Pfizer-BioNTech and Moderna), for the fully vaccinated (2-dose) and those who received a booster (3-dose) of mRNA vaccines during the Delta and Omicron dominant periods^16^ in the US Veteran population, which includes individuals across the U.S. with underlying health conditions and diverse socio-economic backgrounds.

## METHODS

The study was approved by the institutional review board (IRB) of the Department of Veterans Affairs Medical Center in White River Junction, Vermont, which waived the requirement for informed consent. We followed the Strengthening the Reporting of Observational Studies in Epidemiology reporting guideline.

### Data Source

We used electronic medical records and COVID-19 laboratory test data from the VHA Corporate Data Warehouse (CDW) to define infection irrespective of symptoms. The VHA CDW holds electronic medical records for about 9 million U.S. Veterans who use the VHA’s network of 1,293 health care facilities, including hospitals and outpatient clinics. Using CDC variant tracking data^16^, we presumed Omicron SARS-CoV-2 infections those tested positive during January 2022 and Delta those tested positive in November 2021. Because December 2021 represented an overlap of both variants, we excluded infections occurred during this month. As Medicare claims were available only up to August 2021, we used them to find COVID-19 diagnoses, vaccinations, and hospitalizations prior to the study period to exclude patients with prior history of COVID-19 and to augment our vaccination records. Since this population of Veterans was largely over 65 years old, we supplemented CDW data with available Medicare data as Veterans may use Medicare to provide healthcare services even if we found frequent users of VHA. We used VHA CDW records to determine date of death.

### Study Design

To estimate VE against infection, we conducted a matched test negative case-control study (Table 1). The study population included Veterans ages ≥18 years residing in a US state or Washington, D.C., tested for SARS-CoV-2 by PCR (98%) or antigen (2%) testing at a VHA facility during the study period (November 2021 (Delta predominance) or January 2022 (Omicron predominance). We required patients to have enrolment in the VHA and at least one inpatient/outpatient encounter at a VHA facility during the 2 years before the study period and be tested within the VA at an outpatient clinic or emergency room or within one day of hospitalization. As we wanted to study a COVID-19 naïve population, we used VHA records to exclude patients with a known prior history of COVID-19, defined as a VHA diagnosis code or positive antigen/PCR lab test prior to the study period. For those with dual Medicare enrolment, we also excluded patients with any claim with a COVID-19 diagnosis code prior to the study period up until August 2021 (the latest available date for Medicare data). We excluded Johnson & Johnson’s Janssen vaccinees at the date of vaccination, thus excluding their subsequent tests from the analysis. We also censored vaccinees with more than 3 doses of any vaccine at the date of their fourth vaccination. Veterans who tested positive (regardless of symptoms) were considered cases, and those who tested negative, controls. For patients with multiple tests, those negative tests within 10 days of a positive test were excluded (positive tests were kept and negative tests were excluded). We matched each case with up to 4 controls based on Health and Human Service (HHS) geographic region and SARS-CoV-2 laboratory tests within 3 weeks of the case specimen collection date as both are measures of local disease burden. For estimated VE against hospitalization and death, we utilized a matched case-control design. Cases were those who tested positive and were hospitalized or died within 30 days of the positive COVID-19 test. Controls were those not hospitalized or who did not die within 30 days of their SARS-CoV-2 tests, irrespective of test results. Any patient with dual Medicare-VHA enrolment who had a claim for COVID-related hospitalization prior to the study period was already excluded, thus ineligible to be a control.

**Table 1.** Baseline Characteristics of Matched Study Subjects

|  | Delta |  |  | Omicron |  |  |
| --- | --- | --- | --- | --- | --- | --- |
|  | Case (4028) | Control (16112) | SMD^ | Case (19281) | Control (75219) | SMD^ |
| Number of mRNA vaccines, No. (%) |  |  |  |  |  |  |
| 0 | 2276 (57) | 3933 (24) | 69.2 | 7443 (39) | 16758 (22) | 36.1 |
| 2 | 1611 (40) | 8105 (50) | 20.8 | 8091 (42) | 27808 (37) | 10.2 |
| 3 | 141 (4) | 4074 (25) | 65.3 | 3699 (19) | 30391 (40) | 47.7 |
| Vaccine manufacturer, No. (%) |  |  |  |  |  |  |
| Moderna | 1011 (25) | 6930 (43) | 38.5 | 5825 (30) | 30704 (41) | 22.3 |
| Pfizer-BioNTech | 741 (18) | 5249 (33) | 33.0 | 6010 (31) | 27782 (37) | 12.2 |
| No Vaccine Recorded | 2276 (57) | 3933 (24) | 69.2 | 7443 (39) | 16710 (22) | 36.2 |
| Month of Test, No. (%) |  |  |  |  |  |  |
| 2021-November | 4028 (100) | 16112 (100) | 0.0 | n/a | n/a | n/a |
| 2022-January | n/a | n/a | n/a | 19281 (100) | 75219 (100) | 0.0 |
| Age, years, No. (%) |  |  |  |  |  |  |
| 18 to 44 | 866 (21) | 103 (1) | 70.5 | 4317 (22) | 3865 (5) | 51.7 |
| 45 to 64 | 1381 (34) | 5790 (36) | 3.5 | 7669 (40) | 27815 (37) | 5.8 |
| 65 to 74 | 1087 (27) | 6308 (39) | 26.1 | 4692 (24) | 25882 (34) | 22.3 |
| 75 to 84 | 531 (13) | 2809 (17) | 11.8 | 2097 (11) | 13179 (18) | 19.1 |
| 85+ | 163 (4) | 1102 (7) | 12.3 | 506 (3) | 4478 (6) | 16.5 |
| Race, No. (%) |  |  |  |  |  |  |
| Black | 473 (12) | 4547 (28) | 42.1 | 5058 (26) | 22813 (30) | 9.1 |
| Hispanic | 196 (5) | 580 (4) | 6.3 | 1182 (6) | 4229 (6) | 2.2 |
| White | 3099 (77) | 9837 (61) | 34.9 | 11672 (61) | 42405 (56) | 8.5 |
| other | 260 (6) | 1148 (7) | 2.7 | 1369 (7) | 5772 (8) | 2.2 |
| Sex, No. (%) |  |  |  |  |  |  |
| Female | 409 (10) | 1591 (10) | 0.9 | 2643 (14) | 8738 (12) | 6.3 |
| Male | 3619 (90) | 14521 (90) | 0.9 | 16638 (86) | 66481 (88) | 6.3 |
| Quan's CCI |  |  |  |  |  |  |
| Mean (SD) | 1 (1) | 2 (2) | 45.4 | 1 (2) | 2 (2) | 31.9 |
| Median (Q1-Q3) | 0 (0-1) | 1 (0-2) |  | 0 (0-1) | 1 (0-2) |  |
| Abbreviations: Quan's CCI, Charlson comorbidity index, Quan's version<sup>26</sup><br>See Supplemental Table 1 (Young-Xu et al.)<sup>1</sup> for definitions of variables in this table<br>^Standardized mean difference of 10 or greater was used to identify imbalance between cases and controls<sup>27</sup> |  |  |  |  |  |  |

### Exposure

We defined 2-dose vaccination as receipt of two doses of an mRNA COVID-19 vaccine, and booster vaccination as receipt of a third doses of an mRNA vaccine (about 95% of mRNA 3^rd^ doses occurred approximately six months after a 2^nd^ dose). We designated exposure to the two-dose regimen or a booster from 14 days following vaccination, excluding events in days 0-13. We designated patients as unvaccinated if they had no record of vaccination.

### Statistical Analysis

We used conditional logistic regression to calculate odds ratios (ORs) with 95% confidence intervals (CI) for the association between positive SARS-CoV-2 testing and 2 or 3 doses of mRNA COVID-19 vaccine during Delta and Omicron periods. In the model, we included number of mRNA vaccine doses (2 or 3) as the primary explanatory variable, an indicator variable for SARS-CoV-2 presumed variant (Delta or Omicron) based on test date, and their interaction terms. We estimated the association between SARS-CoV-2 infections and number of mRNA vaccine doses by the OR, adjusting for age (continuous), race, rurality, VHA benefits priority, and comorbid conditions (cancer, congestive heart failure, hypertension, immunocompromising conditions, obesity, and diabetes). Confounders were determined based on prior studies^1,2^ and known factors associated with SARS-CoV-2. We also used conditional logistic regression to analyze VE against hospitalizations and death.

Severity of disease was determined by average length of stay (LOS) in days between admission date and discharge date for overall COVID-19-related hospitalizations and average LOS in an intensive care unit (ICU) during a hospitalization related to COVID-19.

All tests were two-tailed, and we chose 0.05 as the level of statistical significance. We performed data analysis using SAS 9.4 (SAS Institute, Cary, North Carolina).

## RESULTS

### Study Population

In November 2021 (Delta period) there were 4,539 positive SARS-CoV-2 tests (cases), and 35,166 negative tests (controls). After matching, 4,028 cases and 16,112 controls remained (see Table 1 for baseline characteristics); 2,276 (57%) cases and 3,933 (24%) controls were unvaccinated at the time of testing. In January 2022 (Omicron period) there were 22,226 positive tests (cases) and 219,209 negative tests (controls). After matching, 19,281 cases and 75,219 controls remained. At the time of testing, 7,443 (39%) cases and 16,710 (22%) controls remained unvaccinated. During both periods, cases tended to be non-Hispanic white, have a lower CCI score and younger than controls. Patients included in this study were largely 65 years or older (62834, 55%), male (101259, 88%), and non-Hispanic white (66986, 58%).

### Vaccine Effectiveness against infection

For those with 2-dose mRNA vaccine, VE was 55% (95% CI, 51 to 58) for Delta and 7% (95% CI, 3 to 10) for Omicron. VE for those who received an mRNA vaccine booster was 90% (95% CI, 88 to 92) in the Delta period – 64% higher than the 2-dose VE, and 59% (95% CI, 57 to 61) in the Omicron period – 7-times higher than the 2-dose VE (Table 2).

**Table 2.** Estimated Vaccine Effectiveness Against Laboratory Confirmed SARS-CoV-2 Infection by Dose and Variant

| Variant | VE (95% CI) | VE Interval (14+ days after vaccination) |
| --- | --- | --- |
| Omicron | 7% (3, 10) | 2<sup>nd</sup> dose |
| Omicron | 59% (57, 61) | 3<sup>rd</sup> dose |
| Delta | 55% (51, 58) | 2<sup>nd</sup> dose |
| Delta | 90% (88, 92) | 3<sup>rd</sup> dose |
| Above numbers exclude Johnson & Johnson's Janssen vaccines as of the date of the Johnson & Johnson's Janssen vaccine. 2<sup>nd</sup> and 3<sup>rd</sup> doses are for mRNA vaccines compared to no vaccination in the indicated period beginning 14 days after vaccination. Tests occurring in 0-13 days after vaccination were excluded. |  |  |
| Cases and controls were matched 1:4 on HHS region and date. The adjusted variables include the following: age (continuous), body mass index, cancer, congestive heart failure, chronic kidney disease, chronic obstructive pulmonary disease, diabetes mellitus, hypertension, immunocompromised, priority level, race/ethnicity, and rurality. |  |  |

### Hospitalization and Death

VE against COVID-related hospitalization for those who received 2-dose mRNA vaccine was 75% (95% CI, 70 to 80) in the Delta period and 44% (95% CI, 26 to 58) in the Omicron period (Table 3). VE against COVID-related hospitalization for those who received an mRNA vaccine booster was 95% (95% CI, 91 to 97) in the Delta period – 27% higher than the 2-dose VE, and 87% (95% CI, 80 to 91) in the Omicron period – twice the 2-dose VE (Table 3). In a sub-analysis restricted to those with a positive COVID-19 test in the study period, we evaluated VE against progression to hospitalization among the infected only, we found that VE for booster against COVID-related hospitalization was 45% (95% CI, -5 to 71) in the Delta period – not statistically different from the 2-dose VE, and 69% (95% CI, 54 to 79) in the Omicron period – almost twice higher than the 2-dose VE (Table S1). Among Omicron-associated hospitalizations, regardless of vaccination status, the length of stay (LOS) averaged 3 days (95% CI, 3 to 4 days), 4 days (95% CI, 3 to 5 days) shorter compared to average Delta hospitalization LOS. Furthermore, 11% (95% CI, 8 to 13%) of hospitalizations during the Delta period resulted in ICU admission; in contrast, 6% (95% CI, 4 to 9%) of COVID-related hospitalizations during the Omicron period resulted in ICU admission (Figure 2).

**Table 3.** Estimated Vaccine Effectiveness Against COVID-19-related Hospitalization and Death by Dose and Variant

| VE Interval (14+ days after vaccination) |  | VE (95% CI) |
| --- | --- | --- |
| Hospitalization |  |  |
| Omicron | 2<sup>nd</sup> dose | 44% (26, 58) |
| Omicron | 3<sup>rd</sup> dose | 87% (80, 91) |
| Delta | 2<sup>nd</sup> dose | 75% (70, 80) |
| Delta | 3<sup>rd</sup> dose | 95% (91, 97) |
| Death |  |  |
| Omicron | 2<sup>nd</sup> dose | 75% (52, 87) |
| Omicron | 3<sup>rd</sup> dose | 94% (85, 98) |
| Delta | 2<sup>nd</sup> dose | 93% (85, 97) |
| Delta | 3<sup>rd</sup> dose | 96% (88, 99) |
| Above numbers exclude Johnson & Johnson's Janssen vaccines as of the date of the Johnson & Johnson's Janssen vaccine. 2<sup>nd</sup> and 3<sup>rd</sup> doses are for mRNA vaccines compared to no vaccination in the indicated period beginning 14 days after vaccination. Tests occurring in 0-13 days after vaccination were excluded. |  |  |
| Cases and controls were matched 1:4 (max) without replacement on HHS and lab test date within three weeks. The adjusted variables include the following: age (continuous), body mass index, cancer, congestive heart failure, chronic kidney disease, chronic obstructive pulmonary disease, diabetes mellitus, hypertension, immunocompromised, priority level, race/ethnicity, and rurality. |  |  |

**Figure 1.**
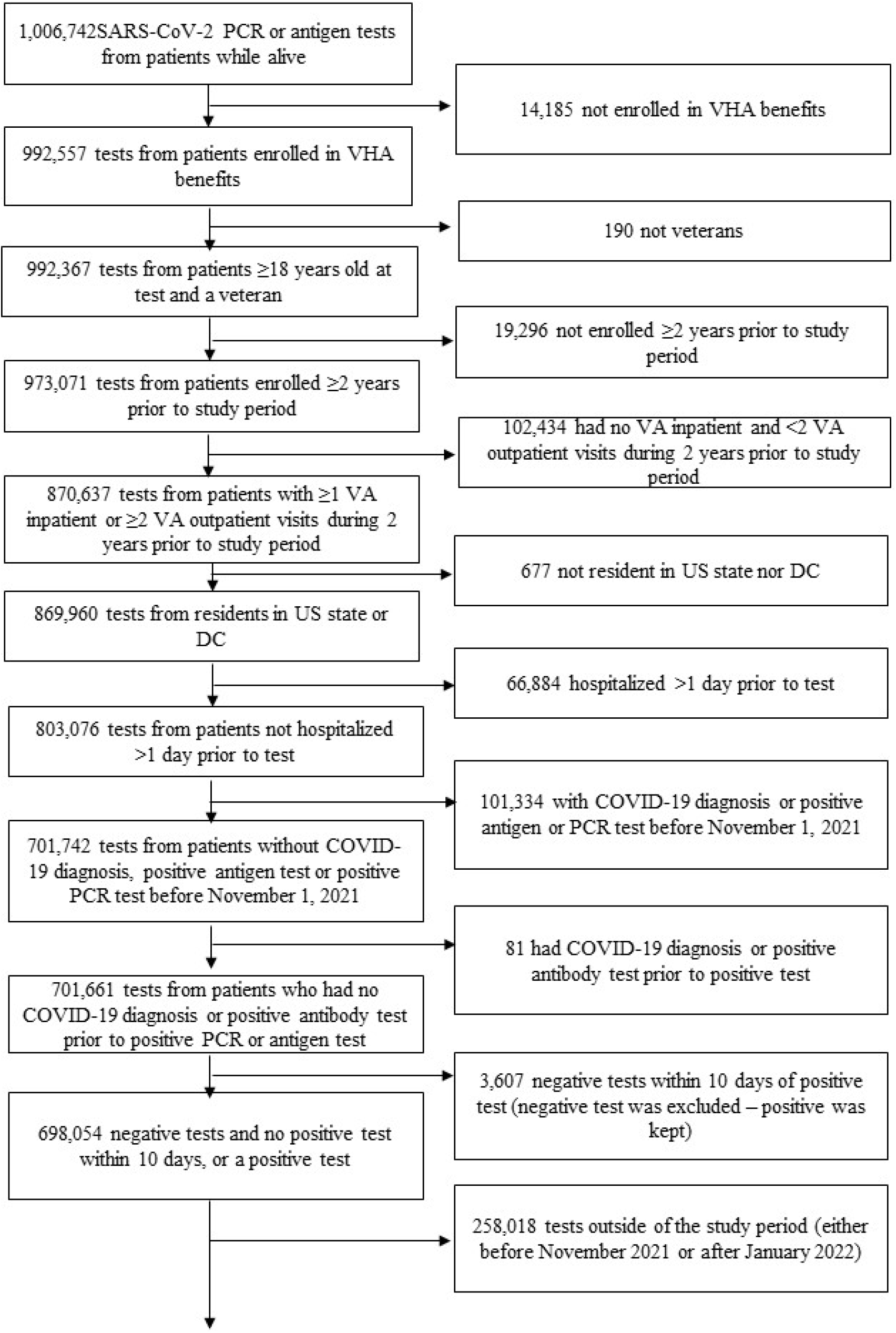
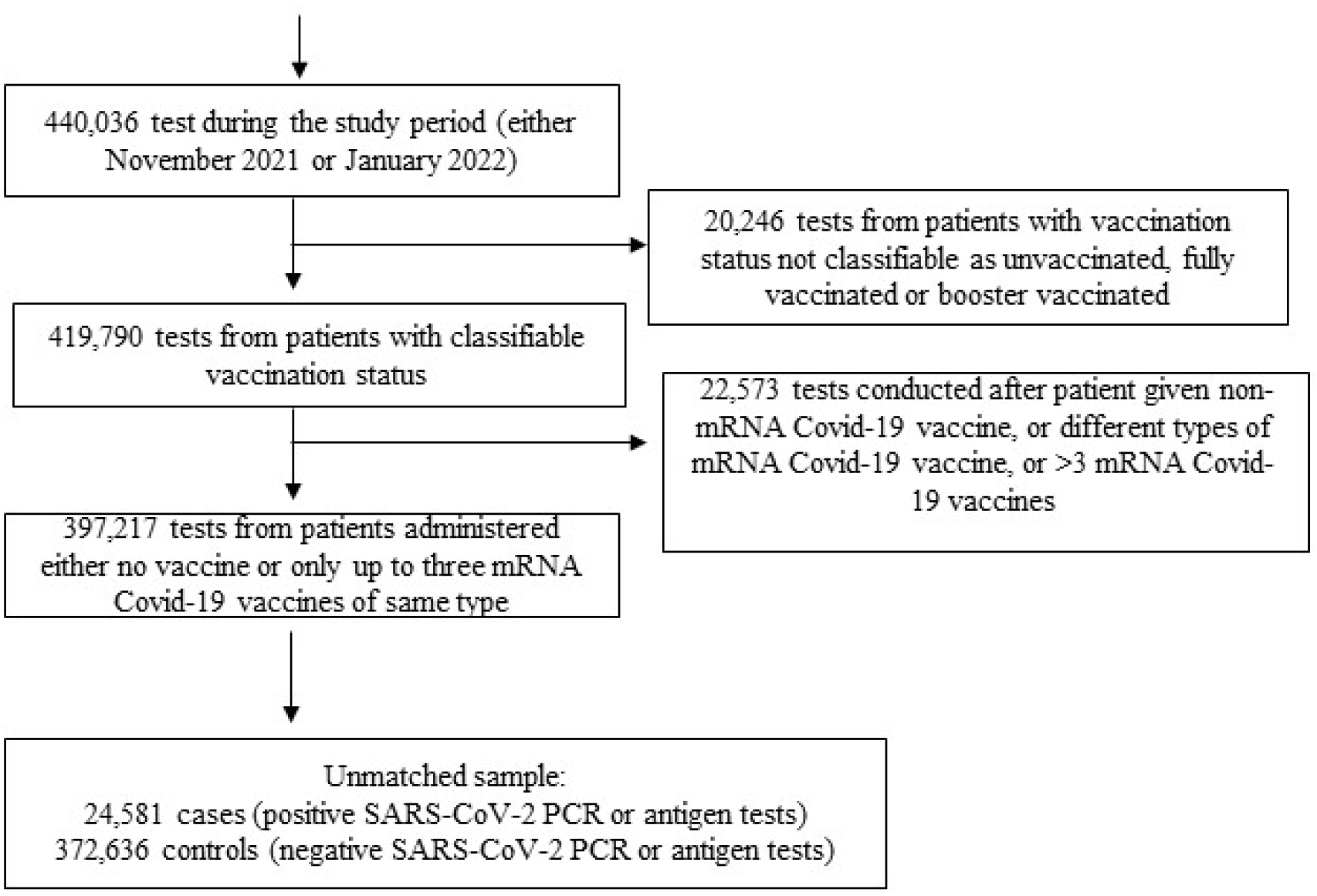
Attrition

**Figure 2.**
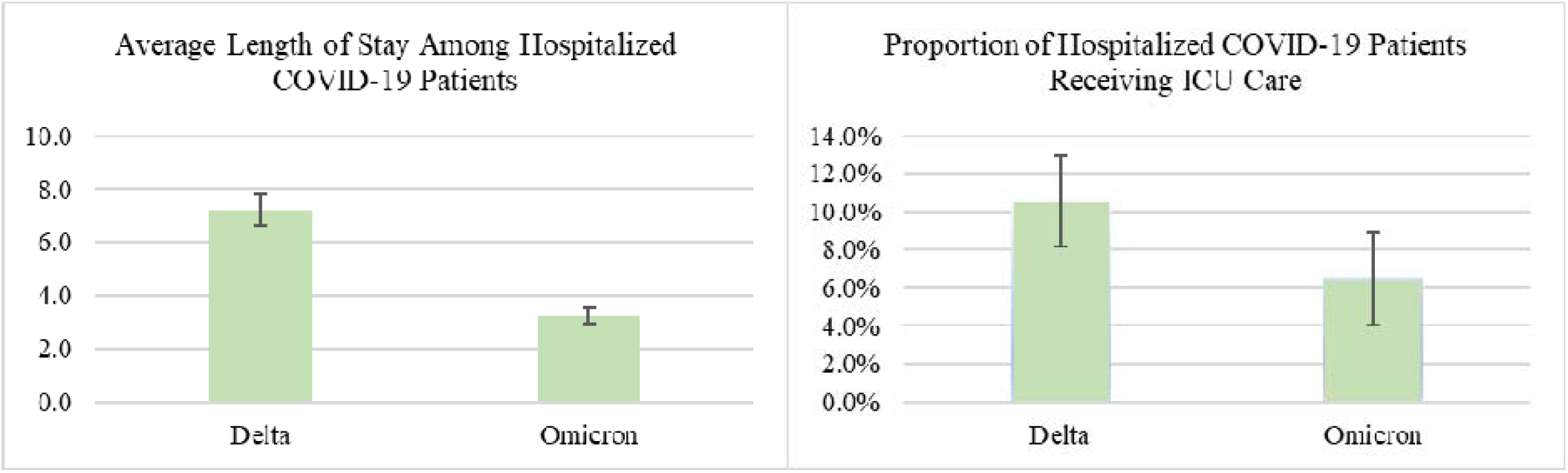
Average Hospitalization Length-of-Stay* and Intensive Care Unit Use During Delta and Omicron Periods * number of days between admission and discharge dates

For those who received 2 doses of mRNA vaccines, VE against death was 93% (95% CI, 85 to 97) during Delta period and 75% (95% CI, 52 to 87) during Omicron period. VE against death for those who received an mRNA vaccine booster was 96% (95% CI, 88 to 99) – no difference from the 2-dose VE - during the Delta period, and 94% (95% CI, 85 to 98) – 25% higher than the 2-dose VE - during the Omicron period.

## DISCUSSION

Our study shows that Veterans who received an mRNA vaccine booster were highly protected against COVID-related outcomes, particularly hospitalization and death, during both the Delta and Omicron periods. These findings largely align with other VE studies of Omicron^11,12,13,14,15,17^. Also, among Veterans tested positive for COVID-19, irrespective of vaccination, those infected during the Omicron period were less likely to be hospitalized and, if hospitalized, had shorter lengths of stay and lower likelihood of ICU admission.

We found that the booster dose significantly increased protection against infection, regardless of symptoms, providing 90% VE against Delta and 59% against Omicron (Table 2) than the 2-dose regimen. These results were generally aligned with published findings. Early results from a study in Scotland showed that booster doses offered a 57% (95% CI, 55 to 60) reduction in risk of symptomatic infection with Omicron compared to a 2-dose regimen.^14^ Similarly, a study from Israel focusing specifically on the Pfizer mRNA vaccine, estimated that a booster vaccination brought vaccine efficacy up to 95% (from around 50% for two-dose vaccination), during a Delta variant period.^18^ Not all studies show a low efficacy for two-dose vaccination during Delta: a U.S. study found a VE of 86.7% (95% CI, 84.3 to 88.7) for two-doses of the Moderna vaccine.^19^ Varying level of waning effectiveness due to different lengths of time since vaccination may explain in part the differences in VE estimates for 2-dose vaccination among the studies. Although we found moderate VE against hospitalization after 2-dose vaccination (Table 3), the booster dose significantly improved VE against hospitalization during the Delta and Omicron periods. For hospitalization, the booster dose increased the VE from 75% to 95% against Delta and from 44% to 87 % against Omicron. Estimated VE of the booster dose against mortality was high during both Delta and Omicron periods: 94% for Omicron and 96% for Delta. These estimates of high VE against mortality resembled those reported elsewhere in the U.S.^20^

A study from South Africa that focused on two-doses of the Pfizer vaccine found an estimated VE against hospitalization of 93% (95% CI, 90 to 94) during the Delta period and 70% (95% CI, 62 to 76) during the Omicron period.^11^ While we found a similar drop in effectiveness from the Delta to the Omicron period, our VE estimates were lower for both periods. Waning protection over time and differences in vaccine coverages and populations may contribute to the disagreement in our findings. For example, studies of hospitalized U.S. Veterans pre-dating the Omicron period showed an estimated VE of 86.8% (95% CI, 80.4-91.1) for full vaccination including mRNA and Johnson & Johnson’s Janssen),^3^ 86.1% (95% CI, 77.7 to 91.3) for 2 doses of Moderna and 75.1% (95% CI, 64.6 to 82.4) for 2 doses of Pfizer-BioNTech.^4^

Our supplementary cases-only analysis showed that the booster vaccination was still highly effective against hospitalization among vaccine breakthrough cases in both the Omicron and Delta periods, with a higher point estimate for the Omicron period (Table S2). This is consistent with studies from the U.K. showing reduced risk of hospitalization among those infected with the Omicron variant compared to the Delta variant.^13,14^ Our results highlight the value of booster doses for protection against infection and, more importantly, against hospitalization and death. They also revealed distinctive features of the two variants, and lower severity of illness for the Omicron variant as compared to the Delta variant.

Strengths of this study include a large, diverse population and near real-time access to medical records. The VHA population offers insight into populations often underrepresented in US studies: racial minorities with a wide range of socio-economic backgrounds. Also, the VHA Veteran population is older (47% above 65), ^25^ sicker, ^23^ and predominantly male (90% male).^25^ Although data on symptomatic versus asymptomatic infection and reasons for SARS-CoV-2 testing were infrequently recorded, in a random sample of Veterans who tested positive and had a detailed record, 26% reported no symptoms (Table S2). Possible misclassification of vaccination status and missing hospitalization records are of concerns because Veterans could go elsewhere for vaccination and hospitalization. For Veterans enrolled in Medicare, we checked records through August 2021 (latest available Medicare data) for COVID-19 vaccination, diagnoses, and related hospitalizations. We also limited the study population to Veterans who routinely sought care at the VHA to minimize misclassification. We assessed patient vaccination status as of November 2021 and January 2022. Other studies, including one of our own, have examined waning mRNA vaccine effectiveness against infection,^2,21,22,23^ so we did not labor further on this topic. Nevertheless, Veterans with underlying conditions and health seeking behaviors may have been vaccinated earlier, thus increased their likelihood of having reduced immunity. Sequencing data were not available for individual SARS-CoV-2 laboratory tests, so we defined variant periods based on estimated variant proportions in the U.S. by the CDC. To reduce misclassification, we excluded December 2021 when Delta and Omicron variants shared dominance, analyzing November 2021 (when Delta was almost 100% dominant) and January 2022 (when Omicron was 90%-100% dominant) separately.^11^ In a future study we plan to use individual sequencing data after they become available in the VHA.

## CONCLUSION

In November 2021, the CDC expanded the booster recommendation to everyone 18 and older 6 months after their second mRNA dose or 2 months after receiving their Johnson & Johnson’s Janssen vaccine.^7^ At that time, 2-dose mRNA vaccine effectiveness among our study population of VHA Veterans were 55%, 75%, and 93% against COVID-19 infection, hospitalization, and death due to the Delta variant, respectively. Although Omicron is highly contagious and more likely to result in breakthrough infections, our findings indicate that mRNA vaccine boosters were very effective against severe COVID-related outcomes during both Delta and Omicron periods. Moreover, the booster vaccination reduced the likelihood of hospitalization among infected patients. Overall, the mRNA vaccine boosters provided a level of protection against Omicron like that of 2-dose against Delta, with VE at 59%, 87%, and 94% against infection, hospitalization, and death, respectively. We can witness the impact of the mRNA vaccine boosters as case numbers continue to drop and the surge of the Omicron variant abates across the United States.

## Data Availability

The data are available upon request to:
Clinical Epidemiology Program
Veterans Affairs Medical Center
215 North Main Street
White River Junction, VT 05009

## Acknowledgements of research support for the study

This project was funded by both the U.S. Food and Drug Administration and the U.S. Department of Veterans Affairs Office of Rural Health.

Yinong Young-Xu had full access to all the data in the study and takes responsibility for the integrity of the data and the accuracy of the data analysis.

## Supplemental Materials Legend

1. Table S1. Estimated Vaccine Effectiveness Against COVID-19-related Hospitalization and Death Among Patients with a Positive Test
2. Table S2. Reasons for Testing and Symptoms

**Table S1.** Estimated Vaccine Effectiveness Against COVID-19-related Hospitalization and Death Among Patients with a Positive Test by Dose and Variant

| VE Interval (14+ days after vaccination) |  | VE (95% CI) |
| --- | --- | --- |
| Hospitalization |  |  |
| Omicron | 2<sup>nd</sup> dose | 24% (-1, 43) |
| Omicron | 3<sup>rd</sup> dose | 69% (54, 79) |
| Delta | 2<sup>nd</sup> dose | 57% (45, 66) |
| Delta | 3<sup>rd</sup> dose | 45% (-5, 71) |
| Death |  |  |
| Omicron | 2<sup>nd</sup> dose | 59% (25, 77) |
| Omicron | 3<sup>rd</sup> dose | 83% (63, 92) |
| Delta | 2<sup>nd</sup> dose | 80% (65, 89) |
| Delta | 3<sup>rd</sup> dose | 60% (24, 87) |
| Above numbers exclude Johnson & Johnson's Janssen vaccines as of the date of the Johnson & Johnson's Janssen vaccine. 2<sup>nd</sup> and 3<sup>rd</sup> doses are for mRNA vaccines compared to no vaccination in the indicated period beginning 14 days after vaccination. Tests occurring in 0-13 days after vaccination were excluded. |  |  |
| Cases and controls were matched 1:4 (max) without replacement on HHS and lab test date within three weeks. The adjusted variables include the following: age (continuous), body mass index, cancer, congestive heart failure, chronic kidney disease, chronic obstructive pulmonary disease, diabetes mellitus, hypertension, immunocompromised, priority level, race/ethnicity, and rurality. |  |  |

**Table S2.** Reasons for Testing and Symptoms in A Random Sample of COVID-19 Tests

|  | Frequency | Percent |
| --- | --- | --- |
| Reason for Testing Recorded (n=1421) |  |  |
| Exposure | 574 | 40% |
| Screening | 847 | 60% |
| Positive Tests (n=606) |  |  |
| Asymptomatic | 160 | 26% |
| Symptomatic | 346 | 57% |
| Unknown | 100 | 17% |
| Total Sample (n=10000) |  |  |
| Records indicating asymptomatic/symptomatic/reason for testing | 1958 | 20% |
| Unknown | 8042 | 80% |
| Exposure: ICD-10-CM code Z20.822; Screening: ICD-10-CM code Z11.52; up to 1 day before and including the date of the SARS-CoV-2 test |  |  |

